# Parents’ intention to vaccinate their 5-11 years old children with the COVID-19 vaccine: rates, predictors and the role of incentives

**DOI:** 10.1101/2021.11.05.21265900

**Authors:** Liora Shmueli

## Abstract

**Background:** On September 20, 2021, Pfizer announced encouraging effectiveness and safety results from their COVID-19 vaccine clinical trials in 5-11 years old children. This study aims to assess parents’ perceptions and intention to vaccinate their 5-11 years old children and to determine the socio-demographic, health-related and behavioral factors, as well as the role of incentives beyond these factors, in predicting this intention.

**Methods:** A cross-sectional representative online survey among parents of children aged 5-11 years in Israel (n=1,012). The survey was carried out between September 23 and October 4, 2021, at a critical time, immediately after Pfizer’s announcement. Two multivariate regressions were performed to determine predictors of parents’ intention to vaccinate their 5-11 years old children against COVID-19 in the coming winter and how soon they intend to do so.

**Results:** Overall, 57% of the participants reported their intention to vaccinate their 5-11 years old children against COVID-19 in the coming winter. This intention was higher for participants over the age of 40. Perceived susceptibility, perceived benefits, perceived barriers, and cues to action, as well as two incentives - vaccine availability and receiving a ‘green pass’ - were all significant predictors of this intention. When asked about how soon they intend to vaccinate their 5-11 years old children, 27% of the participants responded immediately; 26% within three months; and 24% within more than three months. Participants having a family member suffering from a chronic disease as well as those whose children were vaccinated against influenza in the previous winter intend to vaccinate their children sooner. Perceived susceptibility, perceived severity, perceived benefits, perceived barriers, and cues to action, were all found to be significant predictors of this sense of urgency. Similar to the intention to vaccinate children in the coming winter, while vaccine availability and receiving a ‘green pass’ were found to be positive significant predictors of how soon parents intend to vaccinate their children, other incentives such as monetary rewards or monetary penalties were not found to be significant predictors. Parental concerns centered around the safety of the vaccine (64%), fear of severe side effects (60%), and fear that clinical trials and the authorization process were carried out too quickly (56%).

**Conclusions:** This study provides up-to-date information on the rates of the intention of parents to vaccinate their 5-11 years old children, how soon they intend to do so, and the predictors of those intentions, which is essential for health policy makers and healthcare providers for planning vaccination campaigns. Moreover, as vaccine safety and side effects were found to be key parental concerns, it is important to release post-approval safety data regarding the vaccine to the public as soon as such is available. Finally, our findings underscore the important role of vaccine accessibility and receiving a ‘green pass’ over other incentives in promoting parents’ intentions to vaccinate their children.

## 1. Background

The heavy toll of the COVID-19 pandemic on children is becoming increasingly apparent. According to a recent report, children now account for more than one-fourth of COVID-19 weekly reported cases in the US, 1.6%-4.2% of the total cumulated COVID-19 hospitalizations, and 0.00%-0.25% of all COVID-19 deaths (depending on the reporting state) [1]. Many infected children (either symptomatic or asymptomatic) are also experiencing long-term effects, many months after the initial infection, where some studies report rates higher than 50% [2]. Beyond the direct health-related outcomes of an infection, the COVID-19 pandemic has affected children in many other manners, including psychological, educational and economical [3]. Implementing a preventive measure, such as vaccination, to reduce infection among children is therefore critical to reduce severe outcomes and long-term effects in children, as well as to increase community protection.

To date, several vaccines against COVID-19 were authorized by the U.S. Food and Drug Administration agency (FDA). In particular, the Pfizer-BioNTech vaccine was approved for individuals aged >16 years in December 2020 [4] and more recently for children aged 12 to 15 years [5]. While vaccines for younger children were not authorized yet, on September, 20, 2021, Pfizer announced encouraging effectiveness and safety results from its clinical trials of children aged 5 to 11 [6]. However, even when authorized, these vaccines can only be effective if parents desire them.

Despite the availability of COVID-19 vaccines for adults and adolescents, not all eligible individuals choose to vaccinate, in part due to a phenomenon known as vaccine hesitancy. COVID-19 vaccine hesitancy among adults was well-studied. Most of the concerns in this context relate to the safety of the vaccines (e.g., their long term side effects, the speed in which they were developed and authorized, etc.) and their effectiveness (e.g., how long immunity would last, effectiveness against new variants, etc.) [7–10]. However, little is known about parental hesitancy regarding COVID-19 vaccination of their children, and in particular for children aged 5-11, now when such a vaccine is close to authorization.

To actively promote voluntary uptake of COVID-19 vaccines among adults and adolescents, several incentive-based strategies were proposed. Some of the proposed strategies included monetary rewards. For example, in the USA, John Delaney and Robert Litan suggested paying people $1000-1500 for vaccination [11,12]. Another strategy suggested by several governments, including those of Chile, Germany, Italy, the UK, and the USA, was the use of “immunity passports” and “vaccination certificates” [13]. Accordingly, the Israeli Ministry of Health developed an incentives model termed the “green pass”. The ‘green pass’ is an entry permit to facilities and social events, such as hotels, restaurants and concerts, for those who recovered from COVID-19 and for fully vaccinated individuals [12,14]. A previous study found that among incentives, vaccine availability increased the likelihood of getting vaccinated immediately, however incentives such as monetary rewards, monetary penalties or the ‘green pass’ were not found to be significant predictors of getting vaccinated among the adults population [15]. However, to the best of our knowledge, no study thus far examined the role of such incentives in encouraging parents to vaccinate their young children.

As such, the goal of this paper is two-fold: (1) to assess parents’ intention to vaccinate their 5-11 years old children in the coming winter now that a COVID-19 vaccine for that age group is close to authorization and how soon they intend to do so, and (2) to determine the socio-demographic, health-related and behavioral factors, as well as the role of incentives beyond these factors, in predicting these intentions.

## 2. Methods

### Study participants and survey design

A cross-sectional representative online survey among parents of children aged 5-11 years in Israel. The survey was carried out between September 23 and October 4, 2021, immediately after Pfizer announced their results of a clinical trial in children aged 5 to 11 years: the Pfizer-BioNTech’s COVID-19 vaccine was reported to have a favorable safety profile and it appears to generate a robust immune response.

The survey was distributed by Sarid Research Institute for Research Services via an online panel containing a wide audience of potential interviewees who have agreed to participate in surveys from time to time. Eligible participants were required to be (1) 18+ years old and (2) parents of a children aged 5 to 11 years. To compose a representative sample of the Jewish adult population in Israel, respondents were sampled by layers based on geographic area (e.g., large cities and small towns), gender, level of religiosity (secular and orthodox) and socio-economic level.

The questionnaire used in the current study is partly based on a previous questionnaire distributed to the general public in June, 2020, which was prepared after consultation with a panel of experts from public health, including a statistician, a behavioral psychologist and an epidemiologist [16]. After adjusting the questionnaire to the goals of the current study, we refined it by receiving feedback from parents of young children regarding its comprehensibility, usability and time taken to complete the survey. Then, the questionnaire was pre-tested by distributing it to a relatively small group of 100 respondents. After the reliability of the questionnaire was verified (using a Cronbach α test), it was distributed to the rest of the respondents.

### Questionnaire

The questionnaire consisted of the following sections: (1) Socio-demographic predictor variables; (2) health-related predictor variables; (3) Health Belief Model (HBM) predictor variables^1^ and (4) incentives-related predictor variables that could accelerate parents’ readiness to vaccinate their children. Overall, the questionnaire consisted of 40 questions.

### Variables and measurements

The first dependent variable was parents’ intention to vaccinate their 5-11 years old children against COVID-19 in the coming winter, originally measured as a one-item question on a 1-6 scale (1 - not appropriate at all; 6 - very appropriate). This variable was transformed to a binary variable (1 - intend to vaccinate my children; 0 - do not intend to vaccinate my children) so as to simplify the analyses, allowing us to compare individuals who agree to vaccinate their children with those who do not. The second dependent variable was the sense of urgency of parents to vaccinate their 5-11 years old children against COVID-19, measured by 3 categories, namely, intend to vaccinate immediately, intend to vaccinate within 3 months, and intend to vaccinate within more than 3 months.

Independent variables were grouped into four blocks:

1. Socio-demographic predictor variables included: (1) age group; (2) gender; (3) education level; (4) marital status; (5) socio-economic level; (6) periphery level, defined by residential area; (7) religiosity level; and (8) working as a medical staff. The age variable was transformed from numeric to categorical (18-39; 40+) in order to address differences between specific age groups.
2. Health-related predictor variables included: (1) whether the respondent got vaccinated against COVID-19; (2) having a family member suffering from a chronic disease (one or more of the following: heart disease; vascular disease and/or stroke; diabetes mellitus; hypertension; chronic lung disease, including asthma or immune suppression); (3) the existence of past episodes of COVID-19 in the family; (4) the existence of past episodes of hospitalization in the family in the previous year; and (5) whether the children of the respondent got vaccinated against influenza in the previous year.
3. HBM predictor variables included: (1) perceived susceptibility (included two items); (2) perceived severity (included two items); (3) perceived benefits (included three items); (4) perceived barriers (included two item); (5) cues to action (included three items); and (6) health motivation (included one item). Items in HBM were measured on a 1-6 scale (1-not appropriate at all; 6 - very appropriate). Negative items were reverse-scored. Scores for each item were averaged to obtain each of the HBM-independent categories. The Cronbach α internal reliability method revealed the internal consistency of the HBM section to be Cronbach α=0.79 (Table S1).
4. Incentive-related predictor variables included: (1) vaccine availability (“If the vaccine would become accessible and available at school”); (2) monetary rewards; (3) receiving a ‘green pass’ (“If my child would receive a ‘green pass’ that would allow various reliefs (exemption from isolation, entry to places of entertainment etc.)”; and (4) monetary penalties (“If the government would cut my social security benefits or impose another fine in case my child would not get vaccinated”).

### Statistical analyses

Data from the online questionnaires were analyzed using SPSS 26 software, where explicit identifiers were replaced by coded pseudo-identifiers. To test the reliability of HBM measures, Cronbach’s α test was used. To describe characteristics of the study population, the following methods of descriptive statistics were used: frequencies, percentages, averages, and standard deviations.

Relationships between dependent and independent variables were examined by univariate analysis, using either ANOVA or t-tests on independent samples or Chi-squared tests, depending on the characteristics of the variable examined.

Two multivariate regressions were performed. First, to investigate determinants of parents’ intention to vaccinate their children against COVID-19 in the coming winter, a four-step hierarchical binary logistic regression was performed. The intention to vaccinate children served as the dependent variable. In the initial step, only socio-demographic variables that were found to be significant (p<.05) in the univariate analysis were inserted into the regression model as predictors. In the second step, health-related variables that were found to be significant (p<.05) in the univariate analysis were entered as predictors. In the third step, all HBM variables were entered into the model as predictors. In the fourth and final step, four incentive-related variables were entered as predictors (vaccine availability, monetary rewards, monetary penalties and receiving a ‘green pass’).

Second, to estimate the sense of urgency of parents to vaccinate their children against COVID-19, a multinomial logistic regression was estimated. Specifically, I was interested in predicting what would increase the intention of parents to vaccinate their children within 3 months or more than 3 months from the moment the vaccine becomes available, compared to immediately. Only socio-demographic and health-related variables were found to be significant (p<.05) in the univariate analysis were inserted into the regression model as predictors. All HBM and incentive-related variables were entered into the model as predictors.

## 3. Results

### Participant characteristics

Descriptive characteristics of the respondents are provided in Table S2. Overall, 1,012 parents of children aged 5-11 years completed the survey, 51% of whom were female (n=516). Almost half of the parents (n=489) were 18-39 years old. 60% of the respondents (n=625) hold an academic degree and most (89%, n=902) are married. Socio-economic level distributed nearly equally between the three categories (low, medium, and high). 13% of the respondents (n=135) live in peripheral geographical regions. Almost half of the respondents (n=498) were secular. 23% of the respondents (n=231) stated having a family member suffering from a chronic disease. Among respondents who have children aged 12-15 years (42%, n=422), around half (n=214) stated these children were vaccinated against COVID-19.

### Rates of intention to vaccinate children

Overall, 57% of the parents (n=579) intend to vaccinate their 5-11 years old children against COVID-19 in the coming winter. When asked “how soon?”, 27% of the parents (n=270) responded that they intend to vaccinate their children immediately, within less than one month; 26% of them (n=267) answered within one to three months; and 24% (n=242) responded that they would wait (17% would vaccinate within four to 12 months, 7% would wait more than a year). 23% of the parents (n=233) responded that they would not vaccinate their children in this age group at all, and as such were not included in the sense of urgency to get vaccinated analysis.

### Main reasons for the reluctancy to vaccinate children

The main reasons preventing parents from vaccinating their 5-11 years children immediately (n=742) included: concerns about the safety of the vaccine among children (64%), fear of severe side effects of the vaccine (60%), and fear that the vaccine clinical trials and the authorization process were carried out too quickly (56%). Additional reasons included the feeling that COVID-19 isn’t dangerous for children in this age group, so there is no reason to vaccinate them (33%), as well as a concern from a low effectiveness of the vaccine (25%).

### Univariate analyses: the intention to vaccinate children

Results of univariate analyses between socio-demographic and health-related variables and the intention of parents to vaccinate their 5-11 years old children against COVID-19 in the coming winter are reported in Table S2. Demographically, significant differences of this intention exist between men and women (63% vs. 52%, p<0.001, respectively), parents over the age of 40 vs. those younger than 40 (64% vs. 49%, p<0.001, respectively), those with an academic degree vs. those without an academic degree (60% vs. 53%, p<0.05, respectively), and individuals with a higher-than-average income compared to those with a below-average and average income (67% vs. 52%, p<0.001, respectively). Interestingly, parents whose children received the flu vaccine in the previous winter expressed significantly higher willingness to vaccinate their children against COVID-19 in the coming winter than those whose children did not receive the flu vaccine in the previous winter (67% vs. 49%, p<0.001, respectively). Parents who were vaccinated against COVID-19 conveyed greater readiness to vaccinate their 5-11 years old children, compared to those who were not vaccinated (61% vs. 15%, p<0.001, respectively). Among parents who have children aged 12-15 years (n=422), those whose 12-15 years old children were vaccinated against COVID-19, expressed greater intention to vaccinate their 5-11 years old children, compared to those whose 12-15 years old children were not vaccinated (77% vs. 46%, p<0.001, respectively). No significant differences were found between religious denominations, marital status, working as a medical staff, periphery level, having a family member suffering from a chronic disease, past episodes of COVID-19 in the family or past episodes of hospitalization in the family in the previous year.

Results of univariate analyses between HBM and incentive-related variables and the intention to vaccinate 5-11 years old children against COVID-19 in the coming winter are reported in Table S3. Perceived susceptibility, perceived barriers, perceived benefits, cues to action and health motivation, were all found to have a statistically significant effect (p<0.05). No significant effect was found for perceived severity. With respect to incentive-related variables, all four incentives (availability, monetary rewards, monetary penalties and ‘green pass’) were found to have statistically significant effects (p<0.05) on the intention of parents to vaccinate their 5-11 years old children against COVID-19 in the coming winter.

### Univariate analyses: the sense of urgency to vaccinate children

Results of univariate analyses between socio-demographic and health-related variables and the sense of urgency to vaccinate 5-11 years old children against COVID-19 are reported in Table S4. Predictor variables found to have a statistically significant effect (p<0.05) on the sense of urgency are gender, socio-economic level, having a family member suffering from a chronic disease, and whether their children were vaccinated against influenza in the previous winter. Predictor variables not found to have a statistically significant effect include age group, educational level, marital status, periphery level, religiosity level, past episodes of COVID-19 in the family, and past episodes of hospitalization in the family in the previous year.

Results of univariate analyses between HBM variables and incentive-related variables and the sense of urgency to vaccinate 5-11 years old children against COVID-19 are reported in Table S5. The results in this case are completely consistent with those reported in Table S3 for the case of the intention to vaccinate 5-11 years old children against COVID-19 in the coming winter.

### Multivariate analyses: predictors of the intention to vaccinate children

The first regression analysis accounted for an estimated 80% explained variance in intention to vaccinate 5-11 years old children against COVID-19 in the coming winter (Naglekerke’s pseudo *R*^*2*^ = 0.80). All model steps were significant. The most important components of the hierarchical regression were the HBM dimensions, which added 61% to the estimate of the explained variance, on top of the 17% explained by socio-demographic and health-related characteristics. The four incentives added 2% beyond those offered by HBM.

More specifically, according to the final model, among the socio-demographic and health-related variables, only age group was found to be significantly associated with the intention to vaccinate 5-11 years children against COVID-19 in the coming winter, where parents over the age of 40 were 2.45 times likely to vaccinate their children in comparison to parents below the age of 40 (OR=2.45, 95% CI 1.50–4.02). No other socio-demographic nor health-related variables were found to be significant predictors.

Among the HBM variables, perceived susceptibility (OR=2.70, 95% CI 1.97– 3.69), perceived benefits (OR=2.54, 95% CI 1.74–3.69) and cues to action (OR=1.44, 95% CI 1.07–1.94), were all positive significant predictors of the intention to vaccinate 5-11 years old children against COVID-19 in the coming winter. In contrast, perceived barriers (OR=.53, 95% CI.41–.69) was found to be a negative significant predictor. Perceived severity and health motivation were not found to be significant predictors.

Among the incentive-related variables, only vaccine availability (OR=1.69, 95% CI 1.30–2.20) and receiving a ‘green pass’ (OR=1.32, 95% CI 1.04–1.67) were found to be (positive) significant predictors of the intention to vaccinate 5-11 years old children against COVID-19 in the coming winter. Monetary rewards and monetary penalties were not found to be significant predictors.

A complete description of all model steps, goodness of fit indices and regression coefficients are provided in Table 1.

**Table 1.**
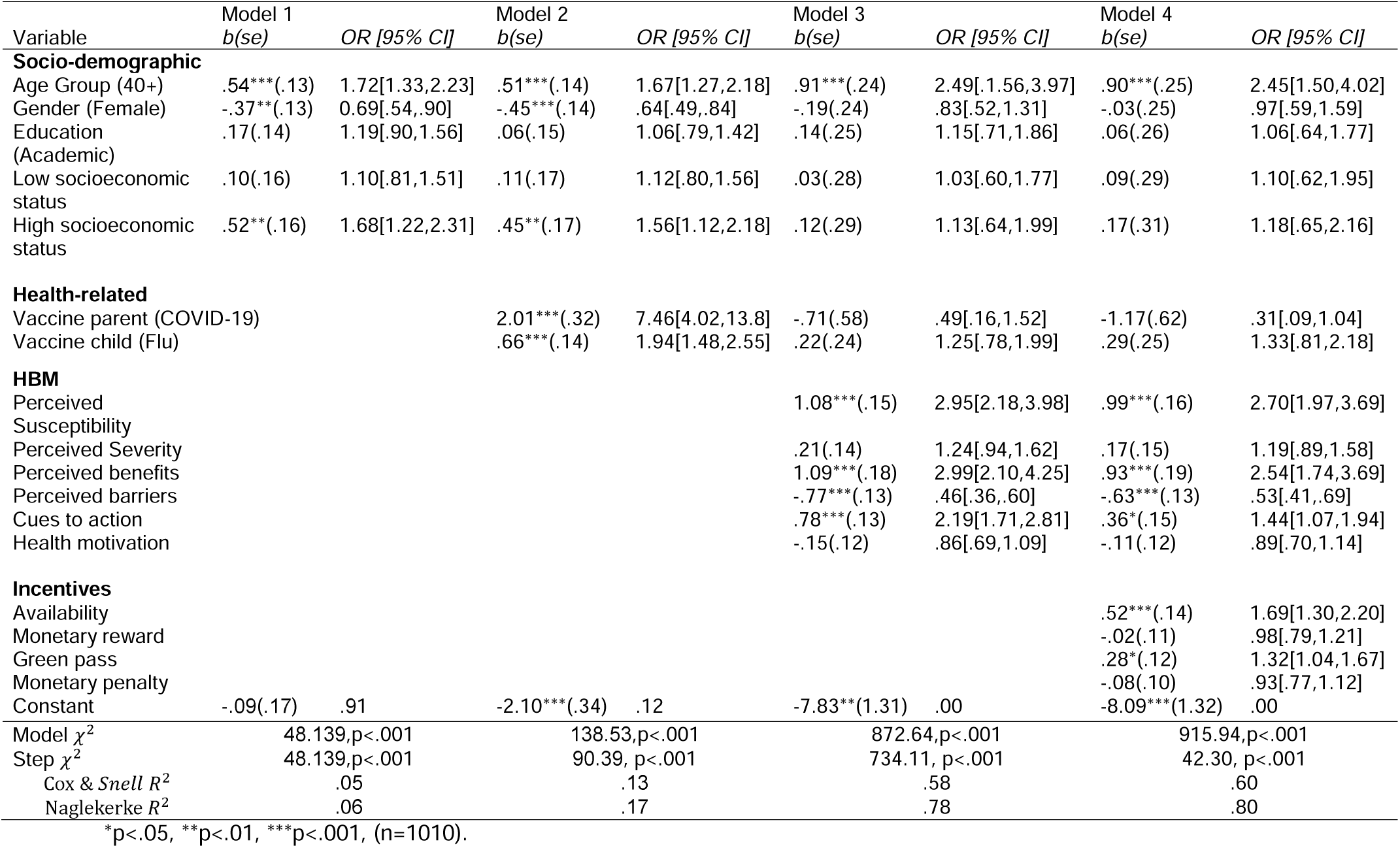
Hierarchical logistic regression analysis - predictors of intention to vaccinate children against COVID-19 (n=1,012)

### Multivariate analyses: predictors of the sense of urgency to vaccinate children

The second regression analyzed the sense of urgency of parents to vaccinate their 5-11 years old children against COVID-19 once the vaccine becomes available for this age group. I was specifically interested in predicting what would increase the intention to vaccinate children in this age group immediately, rather than within 3 months or within more than 3 months from the time the vaccine would become available. The resulting model was found to be significant.

More specifically, according to the resulting model, none of the socio-demographic variables were found to be significantly associated with the sense of urgency to vaccinate 5-11 years old children against COVID-19. From the health-related variables, two were found to be significant predictors. Specifically, having a family member suffering from a chronic disease 0.57 times decreases the intention to vaccinate within 3 months as opposed to immediately (OR=.57, 95% CI.35-.93). The intention to vaccinate within 3 months or within more than 3 months (as opposed to immediately) decreases if their children were vaccinated against influenza in the previous winter (OR=.60, 95% CI.39-.91, OR=.53, 95% CI.30-.92 respectively).

With the exception of health motivation, all HBM variables were found to be significant predictors of the sense of urgency to vaccinate 5-11 years old children against COVID-19. Specifically, for each unit increase in perceived susceptibility, the intention to vaccinate in 3 months rather than immediately decreases .64-fold (OR=.64, 95% CI .47-.86), while the intention to vaccinate within a year rather than immediately decreases .37-fold (OR=.37, 95% CI.25-.55). For each unit increase in perceived severity, the intention to vaccinate in 3 months rather than immediately doesn’t significantly change (OR=.94, 95% CI .76-1.18). Yet, the intention to vaccinate within a year rather than immediately decreases .70-fold (OR=.70, 95% CI.51-.96). Furthermore, for each unit increase in perceived benefits, the intention to vaccinate in 3 months rather than immediately decreases .62-fold (OR=.62, 95% CI.44-.89), while the intention to vaccinate within a year rather than immediately decreases .43-fold (OR=.43, 95% CI.27-.67). Conversely, for each unit increase in perceived barriers, the intention to vaccinate in 3 months rather than immediately increases 1.62-fold (OR=1.62, 95% CI 1.30-2.02), while the intention to vaccinate within a year rather than immediately increases 2.62-fold (OR=2.62, 95% CI 1.92-3.57). Lastly, for each unit increase in cues to action, the intention to vaccinate in 3 months rather than immediately increases 1.33-fold (OR=1.33, 95% CI 1.06-1.68), while the intention to vaccinate within a year rather than immediately doesn’t significantly change (OR=1.27, 95% CI .91-1.77).

Among the incentive-related variables, only vaccine availability and receiving a ‘green pass’ were found to be significant predictors of the sense of urgency to vaccinate 5-11 years old children against COVID-19. Specifically, for each unit increase in vaccine availability, the intention to vaccinate in 3 months decreases 0.7-fold, while the intention to vaccinate within a year decreases 0.48-fold, as compared with the intention to vaccinate immediately (OR=.70, 95% CI .57-.85 and OR=.48, 95% CI .37-.64, respectively). Similarly, for each unit increase in receiving a ‘green pass’, the intention to vaccinate in 3 months decreases 0.72-fold, while the intention to vaccinate within a year decreases 0.50-fold, as compared with the intention to vaccinate immediately (OR=.72, 95% CI .59-.90 and OR=.50, 95% CI .38-.66, respectively). Monetary rewards and monetary penalties were not found to be significant predictors.

A complete description of the model, goodness of fit indices and regression coefficients are presented in Table 2.

**Table 2.** Multinomial logistic regression-predictors of sense of urgency to vaccinate children once COVID-19 vaccine is available for their age group (n=779).

| Variable | Get vaccinated within 3 months |  | Get vaccinated within more than 3 months |  |
| --- | --- | --- | --- | --- |
|  | b(se) | OR (95% CI) | b(se) | OR (95% CI) |
| **Gender** |  |  |  |  |
| Female | .17(.21) | 1.19[.78,1.81] | -.20(.29) | .82[.46,1.45] |
| **Socioeconomic level** |  |  |  |  |
| Low socioeconomic | -.35(.27) | .70[.42,1.18] | -.40(.35) | .67[.34,1.32] |
| High socioeconomic | -.39(.25) | .68[.41,1.11] | -.41(.34) | .67[.34,1.30] |
| **Chronic disease** |  |  |  |  |
| Chronic disease in family | -.56*(.25) | .57[.35,.93] | -.39(.33) | .68[.36,1.30] |
| **Child Flu vaccine** |  |  |  |  |
| Flu vaccinated | -.52*(.21) | .60[.39,.91] | -.64*(.29) | .53[.30,.92] |
| **HBM** |  |  |  |  |
| Perceived susceptibility | -.45**(.15) | .64[.47,.86] | -.99***(.20) | .37[.25,.55] |
| Perceived severity | -.06(.11) | .94[.76,1.18] | -.36*(.16) | .70[.51,.96] |
| Perceived benefits | -.47**(.18) | .62[.44,.89] | -.85***(.23) | .43[.27,.67] |
| Perceived barriers | .48***(.11) | 1.62[1.30,2.02] | .96***(.16) | 2.62[1.92,3.57] |
| Cues to action | .29*(.12) | 1.33[1.06,1.68] | .24(.17) | 1.27[.91,1.77] |
| Health motivation | -.002(.13) | 1.00[.77,1.29] | -.13(.16) | .88[.65,1.21] |
| **Incentives** |  |  |  |  |
| Availability | -.36***(.10) | .70[.57,.85] | -.73***(.14) | .48[.37,.64] |
| Monetary reward | -.05(.07) | .95[.83,1.10] | -.17(.11) | .84[.68,1.04] |
| Green pass | -.32**(.11) | .72[.59,.90] | -.69***(.14) | .50[.38,.66] |
| Monetary penalty | .01(.07) | 1.01[.88,1.17] | .17(.11) | 1.18[.96,1.46] |
| Constant | 5.93***(1.18) |  | 12.15***(1.55) |  |
| $\chi^2(1524) = 3389.024, p < .001$<br>Cox & Snell $R^2 = .55$<br>Naglekerke $R^2 = .62$<br>McFadden $R^2 = .36$ | | | | |

\*p<.05, \*\*p<.01, \*\*\*p<.001

## 4. Discussion and Conclusion

### Discussion

This study assessed parents’ intention to vaccinate their 5-11 years old children and determined the socio-demographic, health-related and behavioral factors, as well as the role of incentives beyond these factors, in predicting this intention.

Only several studies thus far investigated parents’ intention to vaccinate their children, most of them conducted before COVID-19 vaccines were approved for children aged 12-16 years. The present study indicates that more than half of the parents (57%) expressed their intention to vaccinate their children this coming winter if a vaccine against COVID-19 is authorized and becomes available. This finding is consistent with the overall percentage of parents that intend to vaccinate their children (56.8%, averaged over different studies focusing on different age groups of children) shown in a recent systematic review [21]. Several socio-demographic characteristics that we found to be associated with parents’ intention to vaccinate their children are consistent with previous studies. For example, these previous studies found that significantly higher proportions of males [22–24], older parents [21], higher educational level [23,25] and higher income [25,26] exhibited higher intentions to vaccinate children against COVID-19.

We are not aware of any other study investigating how soon parents intend to vaccinate their children against COVID-19. Our study found that only 27% of the parents intend to vaccinate their children immediately. A large proportion of the parents who intended to vaccinate their children preferred to wait a period of time. When these parents were asked about the reasons, the concerns were similar to previous studies and included concerns regarding the vaccines safety, serious side effects and concerns regarding the approval authorization process of the vaccine were carried out too quickly [25,27,28].

Despite this hesitancy of some of the parents’, it is encouraging to see that most of the parental concerns are centered around the safety and effectiveness of the vaccine. Such concerns can be addressed partially once the complete results of the clinical trials, which are expected to show high safety and effectiveness in children, will be released to the public. Moreover, such concerns can be reduced by publishing accessible and transparent information in the media regarding vaccine safety and efficacy as obtained from post-approval data that will be accumulated rapidly as children are vaccinated [27].

In this study, several incentives proposed by health policy makers (i.e. monetary rewards, ‘green pass’, etc.) were added to the predictive model. Among these incentives, receiving a ‘green pass’ and administering the vaccine within the education system were found to increase and accelerate parents’ intention to vaccinate their children. In contrast, neither monetary rewards nor monetary penalties were found to increase parents’ intention to vaccinate their children. This finding is consistent with previous studies demonstrating that monetary incentives do not increase the intention of adults to get vaccinated against COVID-19, mainly since a monetary payment for vaccination is likely to be small and is unlikely to compensate for the risk (perceived or real) of vaccination but only for the inconvenience [29,30]. Moreover, paying people to get vaccinated offends the moral sense of individuals and the community and would have unequal effects on different segments of society [11,31]. These concerns are expected to be magnified when it comes to vaccinating children.

Nevertheless, it is important to recognize this study’s limitations when interpreting the reported results. First, this study was conducted in Israel, and therefore its findings might not necessarily generalize to other countries. Nevertheless, previous studies evaluating vaccination intentions in Israel were found to have consistent trends with those of studies in other countries, and I believe that this study will not be different. Second, our participants were sampled from an online panel so it may miss, inherently, respondents who do not have regular access to the internet. Third, our sample included only the Jewish adult population in Israel, and it did not include the Arab population. Further research should be devoted to reach and include these subpopulations.

### Conclusions

This study provides up-to-date information on the rates of the intention of parents to vaccinate their 5-11 years old children, how soon they intend to do so, and the predictors of those intentions, which is essential for health policy makers and healthcare providers for planning vaccination campaigns. Moreover, as vaccine safety and side effects were found to be key parental concerns, it is important to release post-approval safety data regarding the vaccine to the public as soon as such becomes available. Finally, our findings underscore the important role of vaccine accessibility and receiving a ‘green pass’ over other incentives in promoting parents’ intentions to vaccinate their children.

## Data Availability

The datasets generated during the current study are not publicly available but are available from the corresponding author on a reasonable request.

## Declarations

### Ethics approval and consent to participate

The study was approved by the Ethics Committee for Non-clinical Studies at Bar-Ilan University in Israel. The ethics form was signed by the committee head on September 14, 2021. Consent for participating in the study was obtained digitally through the online questionnaire distributed by Sarid Research Institute. Specifically, at the beginning of the questionnaire, participants were asked whether they agree to participate in the research and whether they were 18 years old or older so as to be included in the study. Only participants who answered these two questions positively were allowed to continue with the questionnaire. Participants were also informed that their participation was voluntary and that they had the right to leave at any time without providing any explanation. Participants in the panel are rewarded for participating in surveys from time to time, according to an estimated time.

### Consent for publication

Not applicable

### Competing interests

The author declares no competing interests.

### Funding

Not applicable

### Authors’ contributions

LS was responsible for study conception and design, data collection and analysis, and writing the manuscript.

### Author information

Liora Shmueli is a lecturer in the Department of Management, Bar-Ilan University.

## Acknowledgements

Not applicable

## Footnotes

1 The HBM is a social psychological model developed to explain and predict health-related behaviors, particularly in regard to the uptake of health services. The HBM has been widely used in the context of vaccination, particularly influenza vaccination [17–19] and more recently COVID-19 [16,20]

## Notes

### Competing Interest Statement

The authors have declared no competing interest.

### Funding Statement

This study did not receive any funding

### Author Declarations

The study was approved by the Ethics Committee for Non-clinical Studies at Bar-Ilan University in Israel. The ethics form was signed by the committee head on September 14, 2021.

## References

[1] Children and COVID-19: State-Level Data Report, (n.d.). http://www.aap.org/en/pages/2019-novel-coronavirus-covid-19-infections/children-and-covid-19-state-level-data-report/ (Accessed October 10, 2021).

[2] Preliminary Evidence on Long COVID in children | medRxiv, (n.d.). https://www.medrxiv.org/content/10.1101/2021.01.23.21250375v1.full (Accessed October 23, 2021).

[3] S. Kamidani, C.A. Rostad, E.J. Anderson, COVID-19 vaccine development: a pediatric perspective, Curr. Opin. Pediatr. 33 (2021) 144–151. https://doi.org/10.1097/MOP.0000000000000978.

[4] K. Dooling, Use of Pfizer-BioNTech COVID-19 Vaccine in Persons Aged ≥16 Years: Recommendations of the Advisory Committee on Immunization Practices — United States, September 2021, MMWR Morb. Mortal. Wkly. Rep. 70 (2021). https://doi.org/10.15585/mmwr.mm7038e2.

[5] O. of the Commissioner, Coronavirus (COVID-19) Update: FDA Authorizes Pfizer-BioNTech COVID-19 Vaccine for Emergency Use in Adolescents in Another Important Action in Fight Against Pandemic, FDA. (2021). https://www.fda.gov/news-events/press-announcements/coronavirus-covid-19-update-fda-authorizes-pfizer-biontech-covid-19-vaccine-emergency-use (Accessed September 29, 2021).

[6] C. Stieg, FDA commissioner Janet Woodcock on Covid vaccine approval for kids: “We’ll do that as quickly as we can,” CNBC. (2021). https://www.cnbc.com/2021/09/21/fda-commissioner-on-covid-vaccine-approval-timeline-for-kids-5-to-11.html (Accessed September 29, 2021).

[7] B. Rosen, R. Waitzberg, A. Israeli, Israel’s rapid rollout of vaccinations for COVID-19, Isr. J. Health Policy Res. 10 (2021) 6. https://doi.org/10.1186/s13584-021-00440-6.

[8] A.A. Dror, N. Eisenbach, S. Taiber, N.G. Morozov, M. Mizrachi, A. Zigron, S. Srouji, E. Sela, Vaccine hesitancy: the next challenge in the fight against COVID-19, Eur. J. Epidemiol. 35 (2020) 775–779. https://doi.org/10.1007/s10654-020-00671-y.

[9] M. Sallam, COVID-19 Vaccine Hesitancy Worldwide: A Concise Systematic Review of Vaccine Acceptance Rates, Vaccines. 9 (2021) 160. https://doi.org/10.3390/vaccines9020160.

[10] J. Wang, X. Lu, X. Lai, Y. Lyu, H. Zhang, Y. Fenghuang, R. Jing, L. Li, W. Yu, H. Fang, The Changing Acceptance of COVID-19 Vaccination in Different Epidemic Phases in China: A Longitudinal Study, Vaccines. 9 (2021) 191. https://doi.org/10.3390/vaccines9030191.

[11] E.A. Largent, F.G. Miller, Problems With Paying People to Be Vaccinated Against COVID-19, JAMA. 325 (2021) 534. https://doi.org/10.1001/jama.2020.27121.

[12] R. Wilf-Miron, V. Myers, M. Saban, Incentivizing Vaccination Uptake: The “Green Pass” Proposal in Israel, JAMA. 325 (2021) 1503–1504. https://doi.org/10.1001/jama.2021.4300.

[13] A.L. Phelan, COVID-19 immunity passports and vaccination certificates: scientific, equitable, and legal challenges, The Lancet. 395 (2020) 1595–1598. https://doi.org/10.1016/S0140-6736(20)31034-5.

[14] What is a Green Pass?, Corona Traffic Light Model Ramzor Website. (n.d.). https://corona.health.gov.il/en/directives/green-pass-info/ (Accessed May 11, 2021).

[15] L. Shmueli, The role of incentives in deciding to receive the available COVID-19 vaccine | medRxiv, (n.d.). https://www.medrxiv.org/content/10.1101/2021.08.11.21261829v1 (Accessed October 11, 2021).

[16] L. Shmueli, Predicting intention to receive COVID-19 vaccine among the general population using the health belief model and the theory of planned behavior model, BMC Public Health. 21 (2021) 804. https://doi.org/10.1186/s12889-021-10816-7.

[17] A. Bish, L. Yardley, A. Nicoll, S. Michie, Factors associated with uptake of vaccination against pandemic influenza: A systematic review, Vaccine. 29 (2011) 6472–6484. https://doi.org/10.1016/j.vaccine.2011.06.107.

[18] T. Kan, J. Zhang, Factors influencing seasonal influenza vaccination behaviour among elderly people: a systematic review, Public Health. 156 (2018) 67–78. https://doi.org/10.1016/j.puhe.2017.12.007.

[19] Rosenstock, Social Learning Theory and the Health Belief Model, (1988). https://journals.sagepub.com/doi/abs/10.1177/109019818801500203?casa_token=8sh1gfDhinIAAAAA:oLFrlO2JbhMy961na4XkKffFCWpFwHuZKJCwpjviTU1uNYjzxxrQ_ak9r7oNlBEj0DPmXrrTXdA4 (Accessed October 15, 2020).

[20] M.C.S. Wong, E.L.Y. Wong, J. Huang, A.W.L. Cheung, K. Law, M.K.C. Chong, R.W.Y. Ng, C.K.C. Lai, S.S. Boon, J.T.F. Lau, Z. Chen, P.K.S. Chan, Acceptance of the COVID-19 vaccine based on the health belief model: A population-based survey in Hong Kong, Vaccine. 39 (2021) 1148–1156. https://doi.org/10.1016/j.vaccine.2020.12.083.

[21] P. Galanis, I. Vraka, O. Siskou, O. Konstantakopoulou, A. Katsiroumpa, D. Kaitelidou, Willingness and influential factors of parents to vaccinate their children against the COVID-19: a systematic review and meta-analysis, Public and Global Health, 2021. https://doi.org/10.1101/2021.08.25.21262586.

[22] R.D. Goldman, S.R. Marneni, M. Seiler, J.C. Brown, E.J. Klein, C.P. Cotanda, R. Gelernter, T.D. Yan, J. Hoeffe, A.L. Davis, M.A. Griffiths, J.E. Hall, G. Gualco, A. Mater, S. Manzano, G.C. Thompson, S. Ahmed, S. Ali, N. Shimizu, Caregivers’ Willingness to Accept Expedited Vaccine Research During the COVID-19 Pandemic: A Cross-sectional Survey, Clin. Ther. 42 (2020) 2124–2133. https://doi.org/10.1016/j.clinthera.2020.09.012.

[23] C.A. Teasdale, L.N. Borrell, S. Kimball, M.L. Rinke, M. Rane, S.A. Fleary, D. Nash, Plans to vaccinate children for COVID-19: a survey of US parents, 2021. https://doi.org/10.1101/2021.05.12.21256874.

[24] Parental Perspectives on Immunizations: Impact of the COVID-19 Pandemic on Childhood Vaccine Hesitancy | SpringerLink, (n.d.). https://link.springer.com/article/10.1007/s10900-021-01017-9 (Accessed October 8, 2021).

[25] M. Skjefte, M. Ngirbabul, O. Akeju, D. Escudero, S. Hernandez-Diaz, D.F. Wyszynski, J.W. Wu, COVID-19 vaccine acceptance among pregnant women and mothers of young children: results of a survey in 16 countries, Eur. J. Epidemiol. 36 (2021) 197–211. https://doi.org/10.1007/s10654-021-00728-6.

[26] E. Hetherington, S.A. Edwards, S.E. MacDonald, N. Racine, S. Madigan, S. McDonald, S. Tough, SARS-CoV-2 vaccination intentions among mothers of children aged 9 to 12 years: a survey of the All Our Families cohort, CMAJ Open. 9 (2021) E548–E555. https://doi.org/10.9778/cmajo.20200302.

[27] P.G. Szilagyi, M.D. Shah, J.R. Delgado, K. Thomas, N. Vizueta, Y. Cui, S. Vangala, R. Shetgiri, A. Kapteyn, Parents’ Intentions and Perceptions About COVID-19 Vaccination for Their Children: Results From a National Survey, Pediatrics. (2021). https://doi.org/10.1542/peds.2021-052335.

[28] K.M. Ruggiero, J. Wong, C.F. Sweeney, A. Avola, A. Auger, M. Macaluso, P. Reidy, Parents’ Intentions to Vaccinate Their Children Against COVID-19, J. Pediatr. Health Care. 35 (2021) 509–517. https://doi.org/10.1016/j.pedhc.2021.04.005.

[29] P. Sprengholz, S. Eitze, L. Felgendreff, L. Korn, C. Betsch, Money is not everything: experimental evidence that payments do not increase willingness to be vaccinated against COVID-19, J. Med. Ethics. (2021). https://doi.org/10.1136/medethics-2020-107122.

[30] S. Pennings, X. Symons, Persuasion, not coercion or incentivisation, is the best means of promoting COVID-19 vaccination, J. Med. Ethics. (2021) medethics-2020-107076. https://doi.org/10.1136/medethics-2020-107076.

[31] N.S. Jecker, What money can’t buy: an argument against paying people to get vaccinated, J. Med. Ethics. (2021). https://doi.org/10.1136/medethics-2021-107235.

[32] T.P. Velavan, A.J. Pollard, P.G. Kremsner, Herd immunity and vaccination of children for COVID-19, Int. J. Infect. Dis. 98 (2020) 14–15. https://doi.org/10.1016/j.ijid.2020.06.065.

[33] P. Klass, A.J. Ratner, Vaccinating Children against Covid-19 — The Lessons of Measles, N. Engl. J. Med. 384 (2021) 589–591. https://doi.org/10.1056/NEJMp2034765.

[34] 40% of new COVID cases in Israel among Arabs, 40% children - The Jerusalem Post, (n.d.). https://www.jpost.com/israel-news/40-percent-of-new-covid-cases-in-israel-among-arabs-40-percent-children-679871 (Accessed October 10, 2021).

