## Supplementary material for "Parents’ intention to vaccinate their 5-11 years old children with the COVID-19 vaccine: rates, predictors and the role of incentives": Table s1

**Table S1. Items and internal consistency for assessing measures of the HBM**

| Measures | Items | a |
| --- | --- | --- |
| Perceived susceptibility | I believe that if I my children will get vaccinated, the likelihood of them getting infected with COVID-19 will decrease | 0.87 |
|  | I believe that if my children will not get vaccinated, the likelihood of our family and relatives getting infected with COVID-19 will increase |  |
| *Perceived severity | Even if my children will get infected with COVID-19 I do not think it will cause them significant suffering or complications | 0.63 |
|  | Even if my children will get infected with COVID-19, the likelihood of them recovering from the disease is very high |  |
| Perceived Benefits | I believe that the COVID-19 vaccine will be highly effective in preventing significant complications of the disease for my children | 0.89 |
|  | I believe that if my children will get vaccinated against COVID-19, the likelihood that they will miss school days (school / kindergarten) will decrease |  |
|  | I believe that if my children will get vaccinated against COVID-19, the likelihood of us (the parents) losing work days will decrease |  |
| Perceived barriers | Getting vaccinated requires time and loss of a workday | 0.68 |
|  | The COVID-19 vaccine does not produce long-term immunity |  |
|  | I am afraid that the COVID-19 vaccine has serious side effects (such as myocarditis etc.) |  |
| Cues to action | The likelihood of vaccinating my children against COVID-19 will increase if friends and family will express their support in the benefits of the vaccine for children | 0.84 |
|  | The likelihood of vaccinating my children against COVID-19 will increase if my pediatrics will recommend me to vaccinate them |  |
|  | The likelihood of vaccinating my children against COVID-19 will increase if the vaccine will be administered in the educational system |  |
| Health motivation | My children are vaccinated with all routine vaccines | - |

a Cronbach indicates the internal consistency: **HBM** a= 0.79

Items Response scale: 1-6 agreement

* Negative items were reverse scored.
