## Supplementary material for "Parents’ intention to vaccinate their 5-11 years old children with the COVID-19 vaccine: rates, predictors and the role of incentives": Tables s2-s5

**Table S2: Characteristics of respondents by the intention to vaccinate their children against COVID-19**

| **Sociodemographic** | **All subjects**  **(n=1,012)** | | **DO not intend to vaccinate children against COVID-19**  **N=433 (42.8%)** | | **Intend to vaccinate children against COVID-19**  **N=579 (57.2%)** | | $\boldsymbol{\chi}\mathbf{2}$ | **p-value** |
| --- | --- | --- | --- | --- | --- | --- | --- | --- |
|  | N | (%) | N | (%) | N | (%) |  |  |
| **Age group** |  |  |  |  |  |  |  |  |
| 18-39 | 489 | (48.3%) | 248 | (50.7%) | 241 | (49.3%) | $\chi2(2)=24.30$ | <0.001 |
| 40+ | 523 | (51.7%) | 185 | (35.4%) | 338 | (64.4%) |  |  |
| **Gender**  Male | 496 | (49%) | 184 | (37.1%) | 312 | (62.9%) | $\chi2(1)=12.87$ | <0.001 |
| Female | 516 | (51%) | 249 | (48.3%) | 267 | (51.7%) |  |  |
| **Education level**  High school or less | 142 | (14%) | 68 | (47.9%) | 74 | (52.1%) | $\chi2(3)=10.84$ | 0.013 |
| Non-academic | 245 | (24.2%) | 115 | (46.9%) | 130 | (53.1%) |  |  |
| BA | 418 | (41.3%) | 181 | (43.3%) | 237 | (56.7%) |  |  |
| MA or higher | 207 | (20.5%) | 69 | (33.3%) | 138 | (66.7%) |  |  |
| **Personal status**  Single | 32 | (3.2%) | 18 | (56.3%) | 14 | (43.8%) | $\chi2(3)=3.65$ | 0.301 |
| Married | 902 | (89.1%) | 386 | (42.8%) | 516 | (57.2%) |  |  |
| Divorced | 74 | (7.3%) | 27 | (36.5%) | 47 | (63.5%) |  |  |
| Widow | 4 | (0.4%) | 2 | (50%) | 2 | (50%) |  |  |
| **Socio-economic level** | | |  |  |  |  | $\chi2(2)=19.52$ |  |
| Low | 336 | (33.2%) | 160 | (47.6%) | 176 | (52.4%) |  | <0.001 |
| Medium | 345 | (34.1%) | 164 | (47.5%) | 181 | (52.5%) |  |  |
| High | 331 | (32.7%) | 109 | (32.9%) | 222 | (67.1%) |  |  |
| **Peripheral level**  Periphery | 135 | (13.4%) | 58 | (43%) | 77 | (57%) |  | 0.96 |
| Intermediate | 439 | (43.5%) | 186 | (42.4%) | 253 | (57.6%) | $\chi2\left( 2 \right)=.09$ |  |
| Central | 436 | (43.2%) | 189 | (43.3%) | 247 | (56.7%) |  |  |
| **Medical Staff** |  |  |  |  |  |  |  |  |
| No | 963 | (95.2%) | 411 | (42.7%) | 552 | (57.3%) | $\chi2\left( 1 \right)=.09$ | .76 |
| Yes | 49 | (4.8%) | 22 | (44.9%) | 27 | (55.1%) |  |  |
| **Religiosity** |  |  |  |  |  |  |  |  |
| Secular | 498 | (49.5%) | 194 | (39%) | 304 | (61%) | $\chi2\left( 3 \right)=7.32$ | .062 |
| Traditional | 284 | (28.2%) | 138 | (48.6%) | 146 | (51.4%) |  |  |
| Religious | 122 | (12.1%) | 55 | (45.1%) | 67 | (54.9%) |  |  |
| Haredi | 103 | (10.2%) | 42 | (40.8%) | 61 | (59.2%) |  |  |

| **Health related variables** | | **All subjects**  **(n=1,012)** | | | **DO not intend to vaccinate children against COVID-19**  **N=433 (42.8%)** | | | **Intend to vaccinate children against COVID-19**  **N=579 (57.2%)** | | $\boldsymbol{\chi}\mathbf{2}$ | **p-value** |
| --- | --- | --- | --- | --- | --- | --- | --- | --- | --- | --- | --- |
|  | N | | | (%) | N | (%) | N | | (%) |  |  |
| **Chronic Disease**  No chronic disease | | | 781 | (77.2%) | 342 | (43.8%) | 439 | | (56.2%) | $\chi2\left( 1 \right)=1.41$ | .24 |
| Chronic disease | | | 231 | (22.8%) | 91 | (39.4%) | 140 | | (60.6%) |  |  |
| **Past episodes of COVID-19**  No | | | 704 | (70.8%) | 296 | (42%) | 408 | | (58%) | $\chi2(1)=0.17$ | .68 |
| Yes | | | 290 | (29.2%) | 126 | (43.4%) | 164 | | (56.6%) |  |  |
| **Flu vaccine last year (kids)** | | |  |  |  |  |  | |  | $\chi2\left( 1 \right)=34.88$ | <.001 |
| No | | | 539 | (53.3%) | 277 | (51.4%) | 262 | | (48.6%) |  |  |
| Yes | | | 473 | (46.7%) | 156 | (33%) | 317 | | (67%) |  |  |
| **A vaccinated parent to COVID-19** | | |  |  |  |  |  | |  | $\chi2\left( 1 \right)=72.81$ | <.001 |
| No | | | 89 | (8.8%) | 76 | (85.4%) | 13 | | (14.6%) |  |  |
| Yes | | | 921 | (91.2%) | 355 | (38.5%) | 566 | | (61.5%) |  |  |
| **Past episodes of hospitalization** | | | | |  |  |  | |  | $\chi2\left( 1 \right)=.43$ | .51 |
| No | | | 919 | (91.8%) | 393 | (42.8%) | 526 | | (57.2%) |  |  |
| Yes | | | 82 | (8.2%) | 32 | (39%) | 50 | | (61%) |  |  |
| **Kids 12-15 vaccinated to COVID-19 (n=422)** | | | | |  |  |  | |  | $\chi2\left( 1 \right)=42.82$ | <.001 |
| No | | | 208 | (49.3%) | 112 | (53.8%) | 96 | | (46.2%) |  |  |
| Yes | | | 214 | (50.7%) | 49 | (22.9%) | 165 | | (77.1%) |  |  |

Note: Percentages of “Do not Intend to get children vaccinated against COVID-19” and “Intend to get children vaccinated against COVID-19” are calculated as valid % per each row (i.e., each row sums up to 100%, without missing values).

**p*<0.05

**Table S3**: Univariate analyses between HBM, incentives variables and willingness to vaccinate your children against COVID-19

|  | DO not-intend to vaccinate children  (n= 433) | | | Intend to get vaccinate children  (n= 579) | | | | | t-test | | P value (two-tail) |
| --- | --- | --- | --- | --- | --- | --- | --- | --- | --- | --- | --- |
| **Variables** | Mean (SD) | | | Mean (SD) | | | | | |  |  |
| **HBM variables** |  | | | |  |  | |  | |  |  |
| Perceived Susceptibility | 2.77 | (1.20) | 4.98 | | | | (.91) | | -33.03 | | <.001 |
| Perceived Severity | 2.52 | (1.10) | 2.55 | | | | (.96) | | -.48 | | .63 |
| Perceived Benefits | 3.19 | (1.20) | 5.07 | | | | (.81) | | -28.18 | | <.001 |
| Perceived Barriers | 4.98 | (1.01) | 3.78 | | | | (1.00) | | 18.78 | | <.001 |
| Cues to action | 2.46 | (1.14) | 4.14 | | | | (1.07) | | -23.79 | | <.001 |
| Health motivation | 4.86 | (1.41) | 5.37 | | | | (.93) | | -6.60 | | <.001 |
| **Incentives variables** |  |  |  | | | |  | |  | |  |
| Availability | 2.05 | (1.20) | 4.60 | | | | (1.34) | | -31.82 | | <.001 |
| Monetary reward | 1.94 | (1.28) | 3.89 | | | | (1.81) | | -20.02 | | <.001 |
| Green pass | 2.43 | (1.47) | 4.94 | | | | (1.27) | | -28.59 | | <.001 |
| Monetary penalty | 2.22 | (1.40) | 3.94 | | | | (1.72) | | -17.5 | | <.001 |

Note: COVID-19 vaccination intention measured by the item: “I want to vaccinated my children against the COVID-19 virus as soon as a vaccine is available”, on a 1-6 agreement scale

HBM and incentives Items Response scale: 1-6 agreement

**Table S4: Characteristics of respondents by sense of urgency to vaccinate their children against COVID-19**

| **Sociodemographic** | **All subjects** | | **Immediately** | | **Within 3 months** | | **More than 3 months** | | $\boldsymbol{\chi}\mathbf{2}$ | **p-value** |
| --- | --- | --- | --- | --- | --- | --- | --- | --- | --- | --- |
|  | N=779 | (%) | N=270 | (34.7%) | N=267 | (34.3%) | N=242 | (31.1%) |  |  |
| **Age group** |  |  |  |  |  |  |  |  | $\chi2\left( 2 \right)=4.76$ | .09 |
| 18-39 | 348 | (44.7%) | 108 | (31%) | 120 | (34.5%) | 120 | (34.5%) |  |  |
| 40+ | 431 | (55.3%) | 162 | (37.6%) | 147 | )34.1%) | 122 | (28.3%) |  |  |
| **Gender**  Male | 398 | (51.1%) | 157 | (39.4%) | 126 | (31.7%) | 115 | (28.9%) | $\chi2(2)=8.24$ | 0.016 |
| Female | 381 | (48.9%) | 113 | (29.7%) | 141 | (37%) | 127 | (33.3%) |  |  |
| **Educational level**  High school or less | 98 | (12.6%) | 39 | (39.8%) | 33 | (33.7%) | 26 | (26.5%) | $\chi2(6)=3.63$ | 0.726 |
| Non-academic | 182 | (23.4%) | 62 | (34.1%) | 65 | (35.7%) | 55 | (30.2%) |  |  |
| BA | 325 | (41.7%) | 104 | (32%) | 111 | (34.2%) | 110 | (33.8%) |  |  |
| MA or higher | 174 | (22.3%) | 65 | (37.4%) | 58 | (33.3%) | 51 | (29.3%) |  |  |
| **Personal status-partnership** | | |  |  |  |  |  |  | $\chi2\left( 6 \right)=4.17$ | 0.653 |
| Single | 21 | (2.7%) | 6 | (28.6%) | 6 | (28.6%) | 9 | (42.9%) |  |  |
| Married | 696 | (89.3%) | 240 | (34.5%) | 239 | (34.3%) | 696 | (31.2%) |  |  |
| Divorced | 59 | (7.6%) | 22 | (37.3%) | 22 | (37.3%) | 59 | (25.4%) |  |  |
| Widow | 3 | (0.4%) | 2 | (66.7%) | 0 | (0%) | 1 | (33.3%) |  |  |
| **Socioeconomic level** | | |  |  |  |  |  |  | $\chi2(4)=11.31$ | 0.023 |
| Low | 237 | (30.4%) | 81 | (34.2%) | 84 | (35.4%) | 72 | (30.4%) |  |  |
| Middle | 256 | (32.9%) | 71 | (27.7%) | 94 | (36.7%) | 91 | (35.5%) |  |  |
| High | 286 | (36.7%) | 118 | (41.3%) | 89 | (31.1%) | 79 | (27.6%) |  |  |
| **Peripheral level** |  |  |  |  |  |  |  |  | $\chi2\left( 4 \right)=1.36$ | 0.851 |
| Periphery | 108 | (13.9%) | 37 | (34.3%) | 34 | (31.5%) | 37 | (34.3%) |  |  |
| Intermediate | 347 | (44.7%) | 123 | (35.4%) | 116 | (33.4%) | 108 | (31.1%) |  |  |
| Central | 322 | (41.4%) | 109 | (33.9%) | 117 | (36.3%) | 96 | (29.8%) |  |  |
| **Medical Staff** |  |  |  |  |  |  |  |  | $\chi2\left( 2 \right)=5.37$ | .068 |
| No | 747 | (95.9%) | 256 | (34%) | 253 | (33.9%) | 238 | (31.9%) |  |  |
| Yes | 32 | (4.1%) | 14 | (43.8%) | 14 | (43.8%) | 4 | (12.5%) |  |  |
| **Religiosity** |  |  |  |  |  |  |  |  | $\chi2\left( 6 \right)=6.40$ | .38 |
| Secular | 404 | (52%) | 147 | (36.4%) | 134 | (33.2%) | 123 | (30.4%) |  |  |
| Traditional | 208 | (26.8%) | 63 | (30.3%) | 74 | (35.6%) | 71 | (34.1%) |  |  |
| Religious | 93 | (12%) | 30 | (32.3%) | 32 | (34.4%) | 31 | (33.3%) |  |  |
| Haredi | 72 | (9.3%) | 30 | (41.7%) | 27 | (37.5%) | 15 | (20.8%) |  |  |

| **Health related variables** | **All subjects** | | | **Immediately** | | **Within 3 months** | | **More than 3 months** | | $\boldsymbol{\chi}\mathbf{2}$ | **p-value** |
| --- | --- | --- | --- | --- | --- | --- | --- | --- | --- | --- | --- |
|  | | N | (%) | N | (%) | N | (%) | N | (%) |  |  |
| **Chronic Disease**  No chronic disease | | 600 | (77%) | 195 | (32.5%) | 219 | (36.5%) | 186 | (31%) | $\chi2\left( 2 \right)=7.29$ | .026 |
| Chronic disease | | 179 | (23%) | 75 | (41.9%) | 48 | (26.8%) | 56 | (31.3%) |  |  |
| **Past episodes of COVID-19**  No | | 562 | (73.4%) | 192 | (34.2%) | 195 | (34.7%) | 175 | (31.1%) | $\chi2\left( 2 \right)=.47$ | .791 |
| Yes | | 204 | (26.6%) | 75 | (36.8%) | 67 | (32.8%) | 62 | (30.4%) |  |  |
| **Flu vaccine last year (kids)** | | | |  |  |  |  |  |  | $\chi2\left( 2 \right)=21.99$ | <.001 |
| No | | 382 | (49%) | 104 | (27.2%) | 135 | (35.3%) | 143 | (37.4%) |  |  |
| Yes | | 397 | (51%) | 166 | (41.8%) | 132 | (33.2%) | 99 | (24.9%) |  |  |
| **A vaccinated parent to COVID-19** | | | |  |  |  |  |  |  | $\chi2\left( 2 \right)=2.28$ | .32 |
| No | | 17 | (2.2%) | 3 | (17.6%) | 7 | (41.2%) | 7 | (41.2%) |  |  |
| Yes | | 761 | (97.8%) | 267 | (35.1%) | 260 | (34.2%) | 234 | (30.7%) |  |  |
| **Past episodes of hospitalization** | | | |  |  |  |  |  |  | $\chi2\left( 2 \right)=.84$ | .66 |
| No | | 705 | (91.2%) | 246 | (34.9%) | 238 | (33.8%) | 221 | (31.3%) |  |  |
| Yes | | 68 | (8.8%) | 24 | (35.3%) | 26 | (38.2%) | 18 | (26.5%) |  |  |
| **Kids 12-15 vaccinated with COVID-19 (n=422)** | | | |  |  |  |  |  |  | $\chi2\left( 2 \right)=2.21$ | .33 |
| No | | 130 | (39%) | 47 | (36.2%) | 42 | (32.3%) | 41 | (31.5%) |  |  |
| Yes | | 203 | (61%) | 82 | (40.4%) | 72 | (35.5%) | 49 | (24.1%) |  |  |

Note: Percentages of “Do not Intend to get vaccinated against COVID-19” and “Intend to get vaccinated against COVID-19” are calculated as valid % per each row (i.e., each row sums up to 100%, without missing values).

**p*<0.05

**Table S5:** Univariate analyses between HBM, incentives variables and the sense of urgency to vaccinate children against COVID-19

|  | **Immediately**  **(n= 270)** | | **Within 3 months (n= 267)** | | **More than 3 months**  **(n= 242)** | | **F-test** | **P value** |
| --- | --- | --- | --- | --- | --- | --- | --- | --- |
| **Variables** | Mean | (SD) | Mean | (SD) | Mean | (SD) |  |  |
| **HBM variables** |  |  |  |  |  |  |  |  |
| Perceived Susceptibility | 5.34 | (.79) | 4.66 | (.91) | 3.62 | (1.03) | 226.67 | <.001 |
| Perceived Severity | 2.50 | (1.00) | 2.63 | (.94) | 2.62 | (.92) | 1.47 | .232 |
| Perceived Benefits | 5.39 | (.71) | 4.79 | (.83) | 3.90 | (.95) | 203.86 | <.001 |
| Perceived Barriers | 3.43 | (1.05) | 4.05 | (.86) | 4.63 | (.93) | 103.36 | <.001 |
| Cues to action | 4.23 | (1.18) | 4.08 | (.93) | 3.27 | (1.04) | 60.27 | <.001 |
| Health motivation | 5.54 | (.81) | 5.29 | (.95) | 4.99 | (1.17) | 20.23 | <.001 |
| **Incentives variables** |  |  |  |  |  |  |  |  |
| Availability | 5.11 | (1.21) | 4.25 | (1.27) | 2.82 | (1.25) | 220.73 | <.001 |
| Monetary reward | 4.33 | (1.84) | 3.65 | (1.70) | 2.45 | (1.43) | 82.36 | <.001 |
| Green pass | 5.42 | (1.04) | 4.67 | (1.30) | 3.24 | (1.36) | 201.30 | <.001 |
| Monetary penalty | 4.30 | (1.80) | 3.68 | (1.59) | 2.86 | (1.45) | 50.32 | <.001 |

Note: the sense of urgency to receive COVID-19 vaccine measured by the item: "now as the vaccine is available, how soon will you get you children vaccinated? Immediately, within 3 months or within a year?".

HBM and incentives Items Response scale: 1-6 agreement
